# Primary Findings of Nationwide Home-based Test to Treat Program for COVID-19 and Influenza

**DOI:** 10.1101/2025.11.13.25339276

**Authors:** Apurv Soni, Biqi Wang, Deogwoon Kim, Caitlin Pretz, Leah Dunkel, Tiffany Israel, Bo Wang, Ben S. Gerber, Shao-Hsien Liu, Gretchen Weaver, Carly Herbert, Nathaniel Hafer, Pragna Patel, Laurel O’Connor, Kathleen Mazor, Kimberly Fisher, Andrew C. Weitz, Home Test to Treat Investigators

**Author notes:** Corresponding Author: Apurv Soni, MD, PhD – Program in Digital Medicine, UMass Chan Medical School, 55 Lake Avenue N, S6-757, Worcester, MA 01655. Funding: This project has been funded in whole or in part with Federal funds from the National Institute of Biomedical Imaging and Bioengineering (NIBIB), National Institutes of Health, Department of Health and Human Services, under Contract No. 75N92022D00010. In addition, the Center for Disease Control (CDC) and the Administration for Strategic Preparedness and Response (ASPR) provided resources for antiviral treatments and over-the-counter tests. The content is solely the responsibility of the authors and does not necessarily represent the official views of the National Institutes of Health.

## Abstract

**Importance:** Timely initiation of antivirals for SARS CoV 2 and influenza depends on rapid diagnosis and access to prescribing. A national home based test to treat service was implemented to address these barriers.

**Objective:** To evaluate reach, use, and timeliness of prescribing in the Home Test to Treat program.

**Design:** Observational cohort analysis of program data from August 15, 2023 through April 17, 2024. Multivariable models assessed factors associated with test kit receipt, telehealth use and timing from symptom onset, antiviral prescribing and timing, and modality of medication fulfillment.

**Setting:** Nationwide, remote program providing free at home tests, on demand telehealth, and oral antivirals with pharmacy pickup or home delivery.

**Participants:** 66,169 adults who enrolled in the program and were eligible for analysis. Participation pathways included 1) proactive testing before symptoms, 2) on demand testing after symptom onset without a diagnosis, and 3) treatment only after a positive test.

**Exposures:** Program pathway and participant characteristics, including insurance status and prior difficulties accessing care.

**Main Outcomes and Measures:** Receipt of test kits, telehealth use and timing from symptom onset, and prescription of guideline directed oral antivirals within 1 day and within 5 days of symptom onset; choice of home delivery versus local pharmacy pickup.

**Results:** Enrollees represented all 50 states and 891 of 896 three-digit ZIP Code areas. Proactive testers comprised 52.6%; 15.0% of the analytical sample reported a positive result. Among positives, 80.7% used the treatment only pathway; 83.2 percent chose telehealth and 76.3% of consults occurred outside local business hours. Antivirals were prescribed to 6,821 enrollees; 59.8% received oral antivirals within 1 day and 92.8% within 5 days of symptom onset. Prescribing within 1 day did not differ notably by age, insurance, race or ethnicity, or prior difficulty accessing care. Of those prescribed, 1,092 enrollees, 16.0%, selected home delivery rather than pharmacy pickup. Home delivery was more common among enrollees reporting more reasons for delaying care, difficulty accessing care, lacking insurance, or enrolled in Medicaid (P < .05).

**Conclusions and Relevance:** Home Test to Treat delivered timely prescribing, including for adults with prior access barriers, by combining home based testing, on demand telehealth, and options for medication delivery across the country.

**Key Points:** *Question:* Does a national home test-to-treat program deliver timely antiviral treatment for COVID-19 and influenza across diverse adults?

*Findings:* Among 66,169 enrollees in all 50 states, 83% of positives used telehealth; 82% received antivirals; 60% were treated within 1 day and 93% within 5 days of symptoms; most visits occurred outside business hours; timeliness did not differ meaningfully by age, insurance, or race/ethnicity once enrolled.

*Meaning:* An integrated home testing and on-demand telehealth pathway achieved rapid, equitable treatment at national scale and provides a practical model to strengthen access and preparedness.

## Introduction

The COVID-19 pandemic exposed significant gaps in healthcare access, particularly in timely testing and treatment for at-risk and vulnerable populations. Early intervention with oral antivirals can prevent severe, adverse outcomes, yet differences in access to care were identified. Individuals from rural areas, racial and ethnic minorities, and vulnerable communities face reduced access to testing and treatment. Non-Hispanic Black and Hispanic patients received outpatient antivirals at substantially lower rates (approximately 30% less) than non-Hispanic White patients. Further, less than a quarter of Medicaid recipients were prescribed oral antivirals during an infection. These findings were persistent despite wide availability in oral antivirals and clear guidelines for indications by the Center for Disease control. The gaps in guideline-directed medical therapy are driven by structural barriers in current public health infrastructure that affect timely recognition of the disease, access to a healthcare provider, assessment of treatment eligibility, and provisioning of treatment. Commonly, the cascade of care requires interaction with different entities: laboratory, clinician, and pharmacists and attrition can emerge at multiple steps.

Emerging care models that co-locate and integrate services promise to overcome the challenges in care continuity. The federal government launched test and treat centers for SARS-CoV-2 as one-stop shop but an estimated 59% of rural residents lived over 60 minutes from the nearest in-person Test-to-Treat site. The site-based test-to-treat programs improved access to some extent, especially in urban centers, individuals in rural and high–social vulnerability index (SVI) areas experienced delays in care. The emergence of home-testing, telehealth, and e-prescriptions can allow for decentralized testing and treatment capabilities. However, programs have focused predominantly on either testing access or antiviral distribution in isolation, without examining the full continuum from diagnosis to treatment. The effectiveness of such a comprehensive home-based test-to-treat approach remains unknown. Potentially, this care model could mitigate known disparities involving the elderly, uninsured, and people of color.

In this study, we evaluate the nationwide Home Test to Treat (HTTT) program, which integrated at-home COVID and influenza testing, telehealth consultations, and medication delivery for COVID-19 and influenza at no cost to the enrollees. The pilot program was designed to address gaps in care for underserved populations through a decentralized model, providing timely diagnosis and treatment via telehealth without in-person visits. By analyzing enrollment patterns, testing outcomes, telehealth uptake, and treatment data, we describe the program’s reach and effectiveness for providing timely guideline directed medical treatment.

## Methods

### Study Design

We conducted a convenience-sampling cohort study of enrollees in the Home Test to Treat (HTTT) program. This national initiative provided at-home COVID-19 and influenza testing, telehealth consultations, and direct medication delivery. Initially conceptualized by a White House task force, HTTT was federally supported by the National Institutes of Health, the Administration for Strategic Preparedness and Response, and the Centers for Disease Control and Prevention. VentureWell, LLC served as the prime contractor and issued a call to coalesce a unique partnership among governmental, non-profit, academic, and private entities. VentureWell contracted UMass Chan Medical School as the research/evaluation partner and eMed, LLC as the technology and implementation partner. Full program details, including pilot phases, are described elsewhere. This report evaluates the main program implementation from August 15, 2023, to April 17, 2024. The study received full ethical approval from the Western Copernicus Group Institutional Review Board (Protocol # 1354231).

### Participants

Individuals U.S. could voluntarily enroll via the program website (www.test2treat.org) or by calling the program hotline (Figure 1). Program enrollment was available to adults aged 18 or older in all 50 U.S. states, the District of Columbia, Puerto Rico, and the U.S. Virgin Islands. Enrollees were classified in one of three pathways: (1) **Proactive test and treat users**: those who enrolled in advance without current symptoms or known infection; (2) **On-demand test and treat users**: individuals who enrolled to request over-the-counter test kits from the program and potential treatment after onset of respiratory symptoms; or (3) **On-demand telehealth users**: individuals who were already COVID-19 or flu positive and enrolled to request telehealth and treatment. (Figure 1) social media, community networks, word of mouth, and planned programmatic press releases were used to spread awareness about the program (Figure 2).

**Figure 1:**
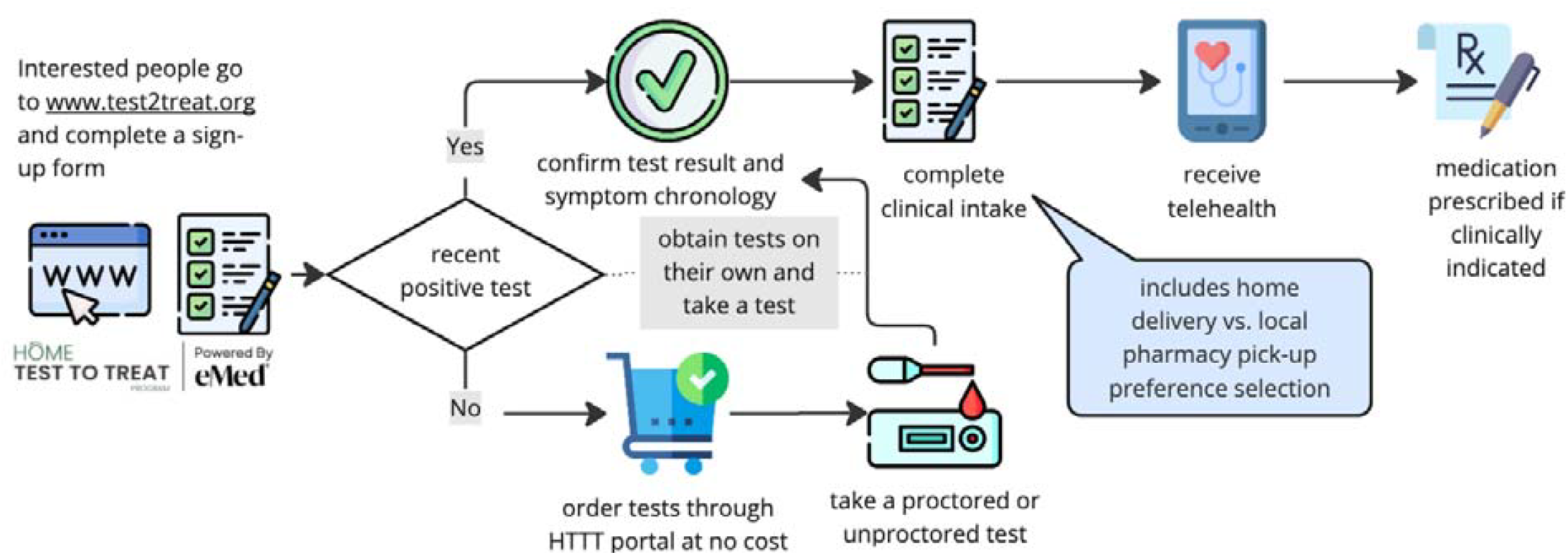
Participant experience in the federal Home-based Test to Treat Program (HTTT)

**Figure 2:**
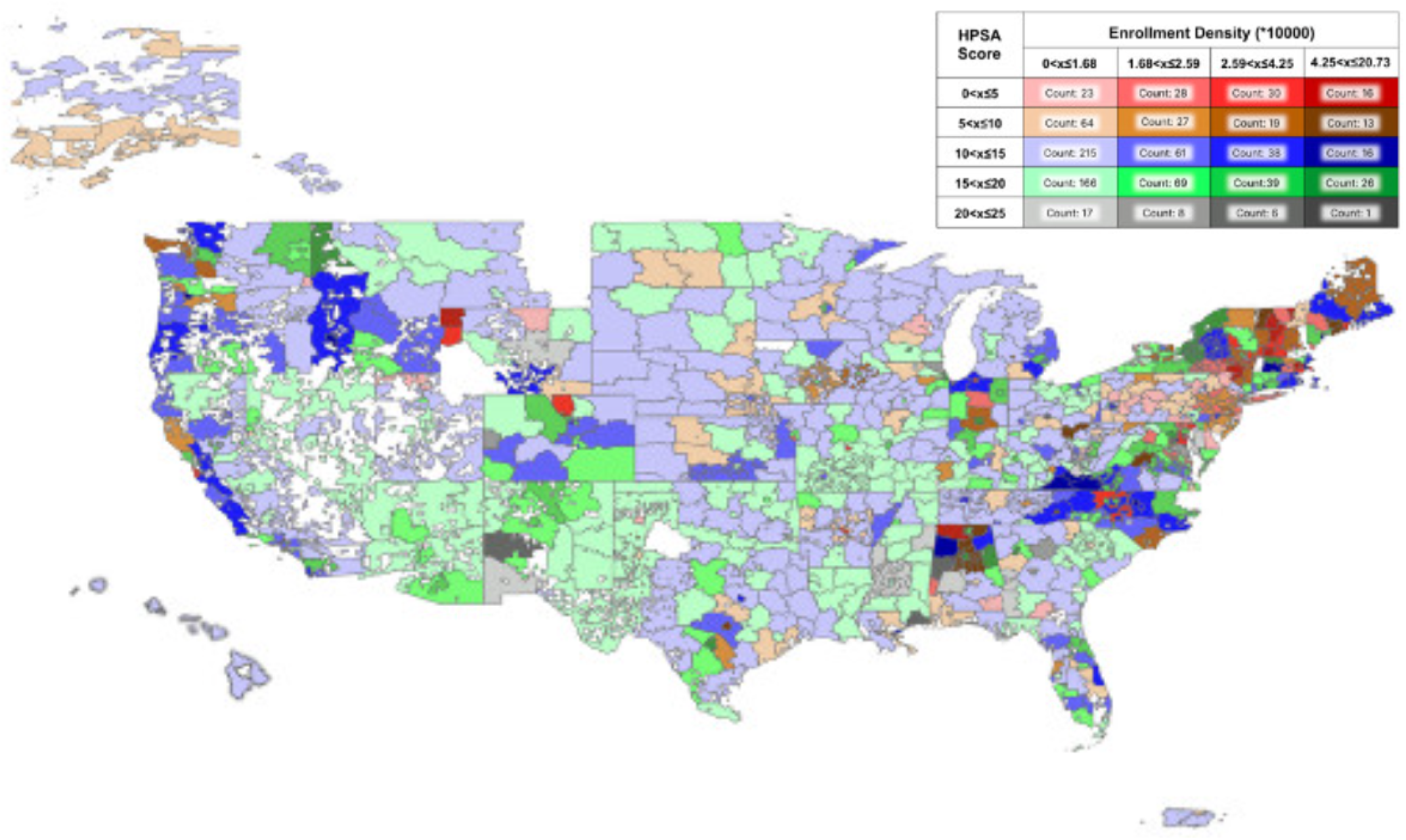
Geospatial coverage of enrollment density in the HTTT program with Health Professional Shortage Area overlay

### Data Collection

#### Data Sources

All data were collected via the HTTT program’s digital platform operated by eMed. Platform metadata provided timestamps for test kit orders, test results reporting, telehealth consultations, and medication prescription. The Health Professional Shortage Area (HPSA) score was obtained from Health Resources and Services Administration (www.data.hrsa.gov).

#### Measures

Upon enrollment, individuals completed a questionnaire on demographics (age, gender, race/ethnicity), recent COVID-19 or flu symptoms or test results, chronic conditions, insurance status, testing and care options, and how they heard about the program. Difficulty in accessing routine clinical care outside of the program was assessed by asking, “If you needed a medical visit tomorrow, how confident are you that you could get it?” Responses on a 4-point Likert scale were dichotomized to “No” (not at all or a little confident) vs. “Yes” (somewhat or very confident). Enrollees were also asked if, in the past year, they had delayed medical care due to limited provider availability, concerns about cost, lack of transportation, or inability to take time off; each reason was recorded as a yes/no response. Primary outcomes were related to treatment provision: (1) the proportion of enrollees who tested positive and received a telehealth consultation, and (2) the proportion of telehealth users who were prescribed an oral antiviral. Secondary outcomes focused on timeliness and access: (1) the proportion of telehealth consultations that began during regular local business hours (9 AM–5 PM weekdays for local time); (4) time from symptom onset to medication prescription; (5) the proportion of enrollees receiving a prescription within 1 and 5 days of symptom onset; and (6) the proportion of enrollees who received medication via home delivery (versus local pharmacy pickup). In the rare event that an enrollee had distinct episodes of positive test results, telehealth encounters, or prescriptions, only the first instance was considered.

### Statistical Analysis

Descriptive statistics were calculated to summarize participant demographics and outcome distributions. Chi-square and ANOVA tests compared differences between the three enrolled groups (proactive HTTT users, on-demand HTTT users, on-demand telehealth seekers) for categorical and continuous variables respectively. Multivariable logistic regression assessed the association between participant characteristics (e.g., race, insurance status, age, gender) and key outcomes such as receipt of telehealth services and timely medication delivery. Adjusted odds ratios (aORs) and 95% confidence intervals (CIs) were reported for associations. All statistical analyses were conducted using R 4.3.3. Significance level was 0.05 (two-sided).

## Results

Of 104,280 individuals who started enrollment process, 80,783 (77.5%) were eligible and completed enrollment (Figure 2). Among those, 14,704 (18.2%) reported a recent positive SARS CoV-2 or influenza test at entry but demonstrated suspicious behavior i.e. only were interested in a research substudy instead of seeking telehealth or treatment. Their data were excluded. The remaining 66,169 enrollees comprised the analytic cohort for this study. More than half of the enrollees had public insurance (Medicare: 31.4%, Medicaid 13.1%) or were uninsured (13.9%), reflecting the program’s emphasis on at-risk and vulnerable population (Table 1). Nearly half of the cohort (44.7%) self-reported lacking confidence in getting needed care within 24 hours outside of the program and more than half had one or more reason to delay healthcare despite a need due to a barrier. Overall, 52.6% enrolled proactively,15.2% enrolled as on-demand test-and-treat users, and the rest (32.2%) were on-demand telehealth seekers. Key differences emerged between the groups: 56.0% of on-demand test-and-treat users, 46.1% of proactive users, and 37.1% of on-demand telehealth seekers indicated difficulty accessing timely care outside of the program. Proactive test and treat users skewed older (38.6% were age ≥65). The on-demand treatment group had a higher proportion of males than the proactive group (39.9% vs. 26.8% respectively). Enrollees who learned of the program via community outreach or friends/family were more likely to enroll as on-demand treatment seekers.

**Table 1:** Key characteristics of participants by group.

| Variable | Overall<br>(n=66,169) | Proactive test<br>and treat users<br>(n=34,830) | On-demand test<br>and treat users<br>(n=10,032) | On-demand<br>telehealth<br>users<br>(n=21,307) |
| --- | --- | --- | --- | --- |
| **Insurance** |  |  |  |  |
| Private | 24,394 (36.9) | 8,997 (25.8) | 1,522 (15.2) | 13,875 (65.1) |
| Medicaid | 8,663 (13.1) | 4,840 (13.9) | 2,448 (24.4) | 1,375 (6.5) |
| Medicare | 20,763 (31.4) | 15,610 (44.8) | 2,829 (28.2) | 2,324 (10.9) |
| No insurance | 9,192 (13.9) | 3,831 (11.0) | 2,772 (27.6) | 2,589 (12.2) |
| Other | 3,157 (4.8) | 1,552 (4.5) | 461 (4.6) | 1,144 (5.4) |
| **Difficulty**<br>**accessing care** | 29,562 (44.7) | 16,053 (46.1) | 5,613 (56.0) | 7,896 (37.1) |
| **Number of**<br>**reasons for**<br>**delaying care** |  |  |  |  |
| 0 | 28,640 (43.3) | 18,458 (53.0) | 3,506 (34.9) | 6,676 (31.3) |
| 1 | 27,429 (41.5) | 11,101 (31.9) | 4,091 (40.8) | 12,237 (57.4) |
| 2 | 7,478 (11.3) | 3,985 (11.4) | 1,711 (17.1) | 1,782 (8.4) |
| 3 | 2,622 (4.0) | 1,286 (3.7) | 724 (7.2) | 612 (2.9) |
| **Age Group** |  |  |  |  |
| 18-45, N (%) | 32,597 (49.3) | 14,198 (40.8) | 5,954 (59.4) | 12,445 (58.4) |
| 46-65, N (%) | 15,961 (24.1) | 7,162 (20.6) | 2,081 (20.7) | 6,718 (31.5) |
| 65+ | 17,595 (26.6) | 13,456 (38.6) | 1,997 (19.9) | 2,142 (10.1) |
| **Gender** |  |  |  |  |
| Female | 45,415 (68.6) | 25,171 (72.3) | 7,510 (74.9) | 12,734 (59.8) |
| Male | 20,207 (30.5) | 9,322 (26.8) | 2,373 (23.7) | 8,512 (39.9) |
| Other | 547 (0.8) | 337 (1.0) | 149 (1.5) | 61 (0.3) |
| **Gender Minority** | 15,613 (23.6) | 9,248 (26.6) | 3,056 (30.5) | 3,309 (15.5) |
| **Race/Ethnicity** |  |  |  |  |
| Hispanic | 5,284 (8.0) | 2,418 (6.9) | 1,129 (11.3) | 1,737 (8.2) |
| Non-Hispanic Black | 7,312 (11.1) | 1,966 (5.6) | 778 (7.8) | 4,568 (21.4) |
| Non-Hispanic White | 45,059 (68.1) | 26,031 (74.7) | 6,578 (65.6) | 12,450 (58.4) |
| Non-Hispanic Other | 8,514 (12.9) | 4,415 (12.7) | 1,547 (15.4) | 2,552 (12.0) |
| **Chronic Condition** | 33,542 (50.7) | 19,884 (57.1) | 6,004 (59.8) | 7,654 (35.9) |
| **Source of referral (not mutually exclusive)** |  |  |  |  |
| Community | 7,818 (11.8) | 1,762 (5.1) | 681 (6.8) | 5,375 (25.2) |
| Social Media | 33,104 (50.0) | 18,592 (53.4) | 4,719 (47.0) | 9,793 (46.0) |
| Friends/Family | 21,464 (32.4) | 9,167 (26.3) | 2,601 (25.9) | 9,696 (45.5) |
| Others | 13,515 (20.4) | 6,887 (19.8) | 2,551 (25.4) | 4,077 (19.1) |
| **HPSA Score**<br>**(mean (SD))** | 13.12 (4.8) | 12.94 (5.00) | 13.15 (4.7) | 13.40 (4.7) |

Difficulty accessing care defined as:
Reasons for delaying care included: limited provider availability, concerns about cost, lack of transportation, or inability to take time off
HPSA: Health Professional Shortage Area Score (HPSA) obtained from data.hrsa.gov

A total of 9,940 enrollees (15.0%) reported a positive COVID-19 or flu test. Across all enrollment pathways, 83.2% of those who tested positive proceeded to receive a telehealth consultation through HTTT. Only 23.7% of these telehealth visits began during regular weekday hours (9 AM–5 PM, local time). Among telehealth users, 82.4% were prescribed an antiviral medication. Overall, 59.8% of enrollees who were given a prescription received it within 1 day of symptom onset, and 92.8% received it within 5 days (Table 2).

**Table 2:**
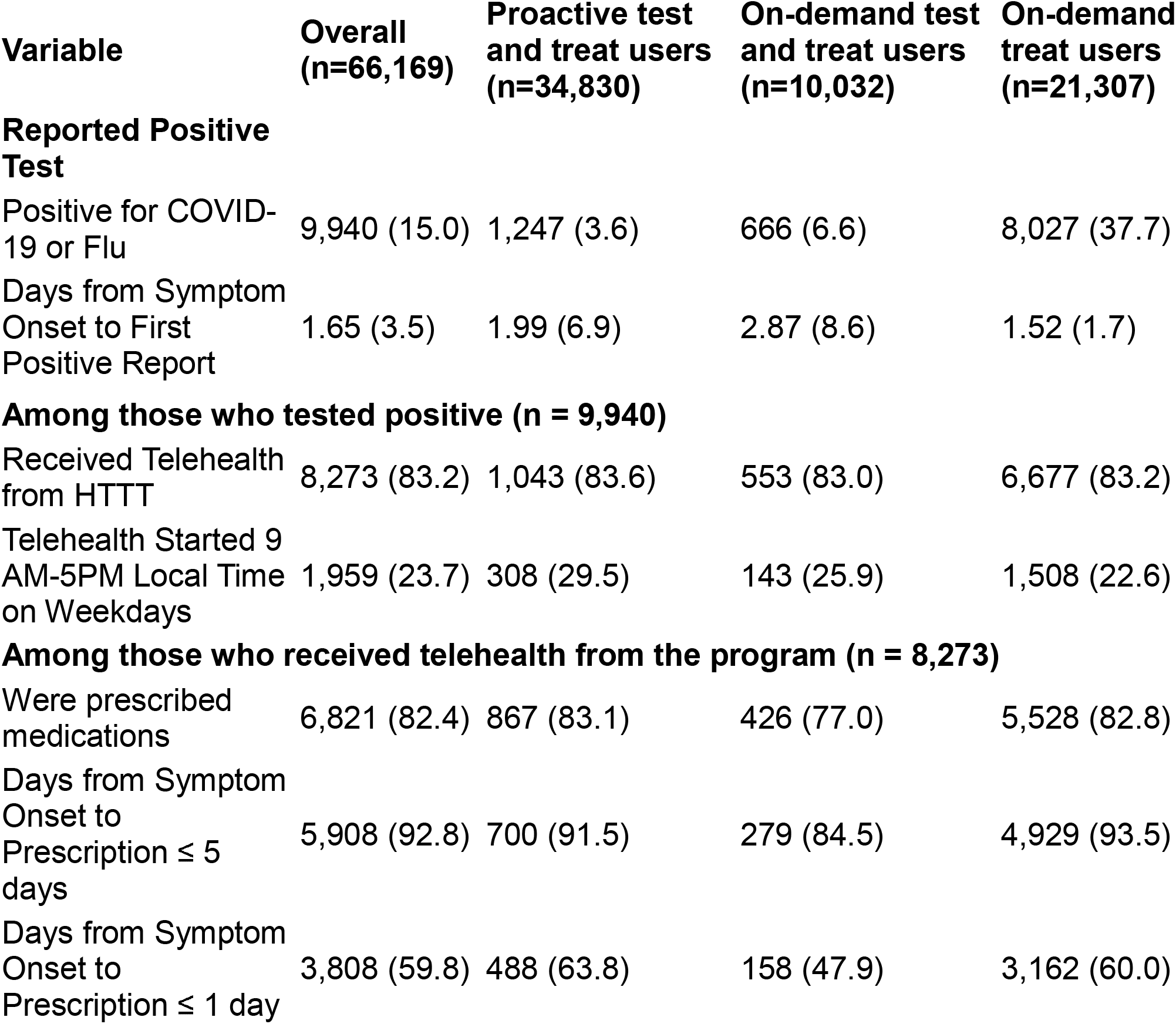
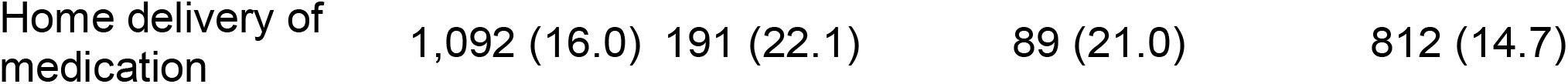
Test and treatment outcomes by group.

| Variable | Overall<br>(n=66,169) | Proactive test<br>and treat users<br>(n=34,830) | On-demand test<br>and treat users<br>(n=10,032) | On-demand<br>treat users<br>(n=21,307) |
| --- | --- | --- | --- | --- |
| **Reported Positive**<br>**Test** |  |  |  |  |
| Positive for COVID-19 or Flu | 9,940 (15.0) | 1,247 (3.6) | 666 (6.6) | 8,027 (37.7) |
| Days from Symptom Onset to First Positive Report | 1.65 (3.5) | 1.99 (6.9) | 2.87 (8.6) | 1.52 (1.7) |
| **Among those who tested positive (n = 9,940)** |  |  |  |  |
| Received Telehealth from HHTT | 8,273 (83.2) | 1,043 (83.6) | 553 (83.0) | 6,677 (83.2) |
| Telehealth Started 9 AM-5PM Local Time on Weekdays | 1,959 (23.7) | 308 (29.5) | 143 (25.9) | 1,508 (22.6) |
| **Among those who received telehealth from the program (n = 8,273)** |  |  |  |  |
| Were prescribed medications | 6,821 (82.4) | 867 (83.1) | 426 (77.0) | 5,528 (82.8) |
| Days from Symptom Onset to Prescription ≤ 5 days | 5,908 (92.8) | 700 (91.5) | 279 (84.5) | 4,929 (93.5) |
| Days from Symptom Onset to Prescription ≤ 1 day | 3,808 (59.8) | 488 (63.8) | 158 (47.9) | 3,162 (60.0) |
| Home delivery of medication | 1,092 (16.0) | 191 (22.1) | 89 (21.0) | 812 (14.7) |

In multivariable adjusted analyses, we observed only limited differences based on participant and regional characteristics. Enrollees reporting difficulty accessing care were more likely to utilize telehealth when positive (aOR 1.35, 95% CI 1.20–1.51). In contrast, older adults (aOR 0.81, 95% CI 0.65–1.00) and Medicare beneficiaries (aOR 0.71, 95% CI 0.58–0.88) were less likely to opt for a telehealth consultation after testing positive. Black enrollees and those with access barriers had higher telehealth uptake, whereas seniors and Medicare-insured enrollees had lower uptake (Figure 3). Among telehealth users, men had higher odds of being prescribed an antiviral treatment (aOR 1.29, 95% CI 1.13–1.48). Conversely, both Medicare beneficiaries (aOR 0.69, 95% CI 0.55–0.88) and Hispanic enrollees (aOR 0.75, 95% CI 0.62–0.91) were less likely to receive an antiviral prescription during their telehealth encounter (Figure 4a). However, there were no meaningful differences observed among antiviral prescription rates within one day of symptom of onset (Figure 4b).

**Figure 3:**
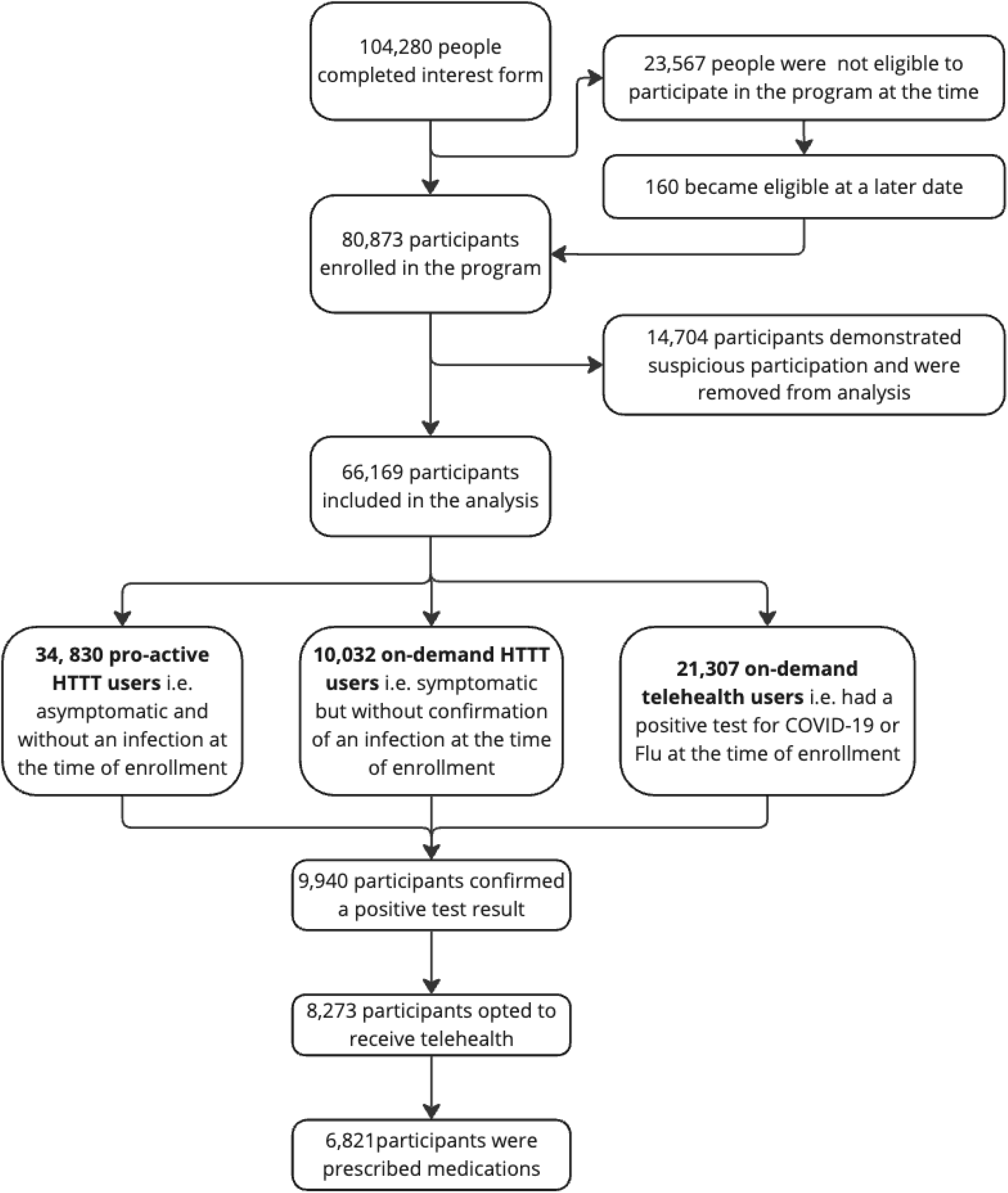
CONSORT diagram of participants in the federal Home-based Test to Treat Program (HTTT) a: After September 13, 2023 enrollment in the program was restricted to people on non-private insurance or without insurance *unless* they had recently tested positive for COVID-19 or Flu. The ineligible participants were placed in a waitlist and allowed to participate if they reporteda positive test result b: suspicious participation refers to participants who enrolled the program after testing positive for COVID-19 and Flu and do not engage in Home-based Test to Treat activities but participated in the research sub-study

**Figure 4a:**
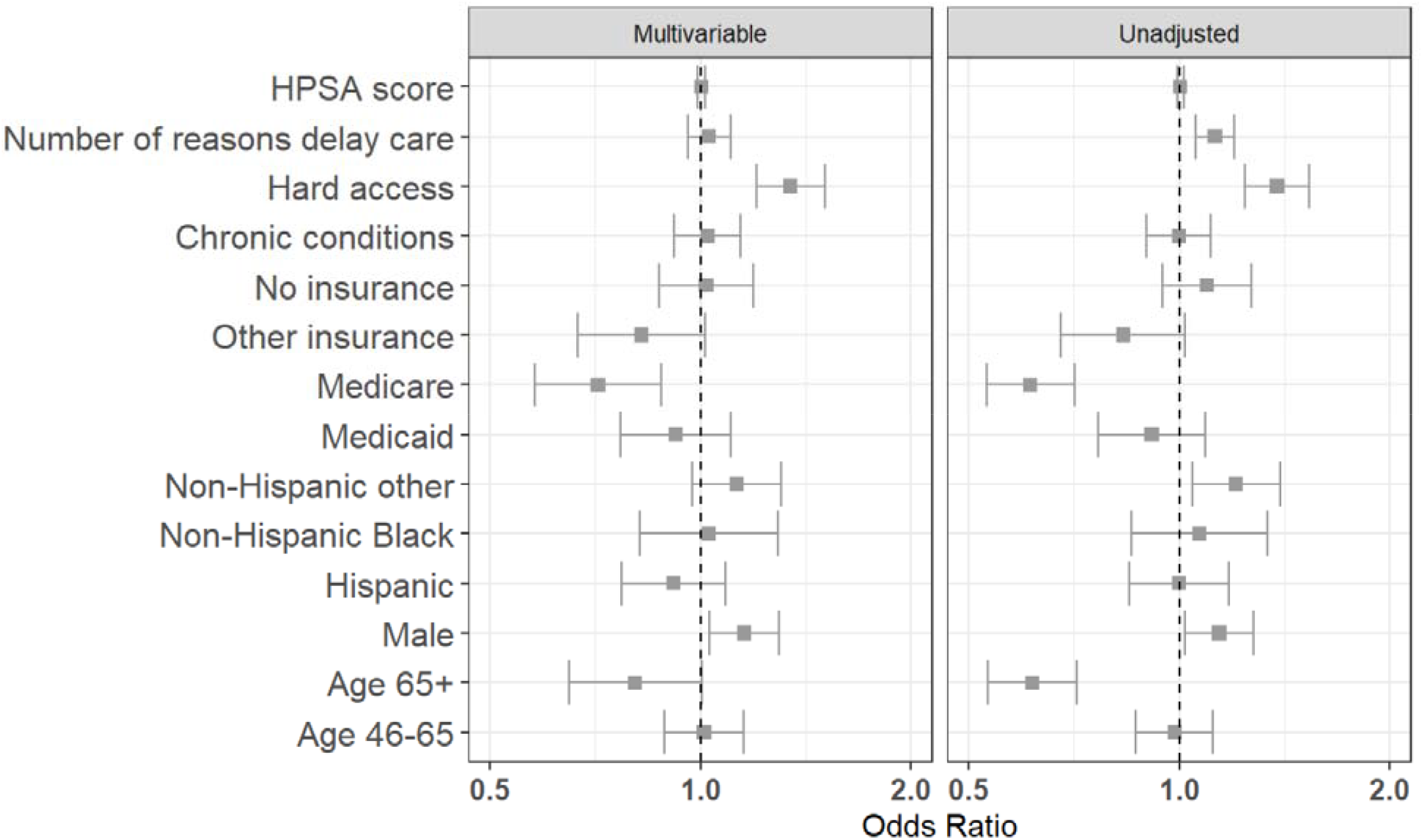
Receive a telehealth consult.

**Figure 4b:**
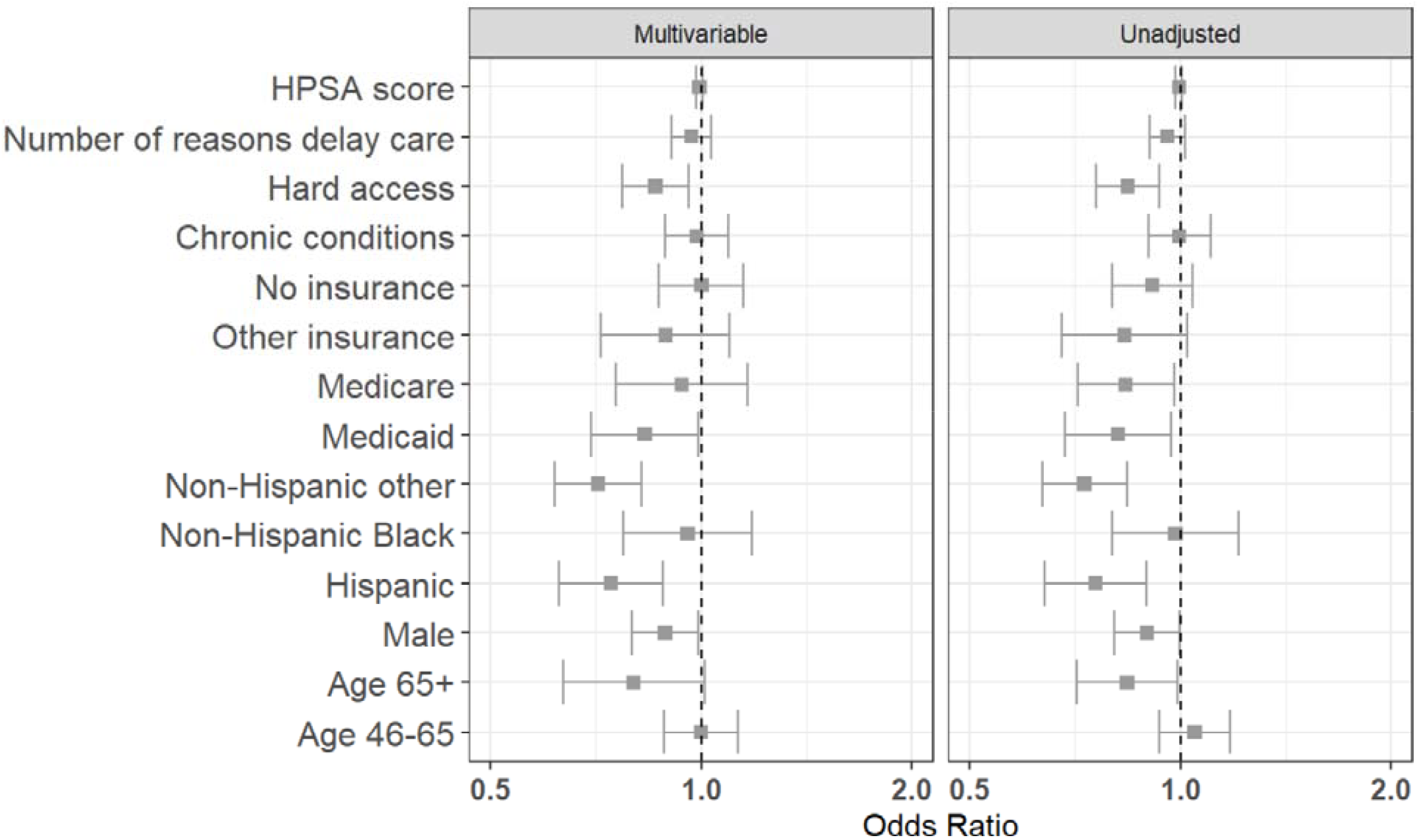
Start of telehealth within business hours.

Although few enrollees chose home delivery of medication (16.0% overall), this option was more commonly utilized by those facing access barriers. Enrollees who reported difficulty accessing care (aOR 1.27, 95% CI 1.09–1.47), who had any recent delays in seeking care (aOR 1.31, 95% CI 1.21–1.42), or who were uninsured (aOR 1.29, 95% CI 1.08–1.53) or enrolled in Medicaid (aOR 1.63, 95% CI 1.32–2.01) had higher odds of opting for mail-order medication delivery rather than local pharmacy pickup (Figure 5).

**Figure 5a:**
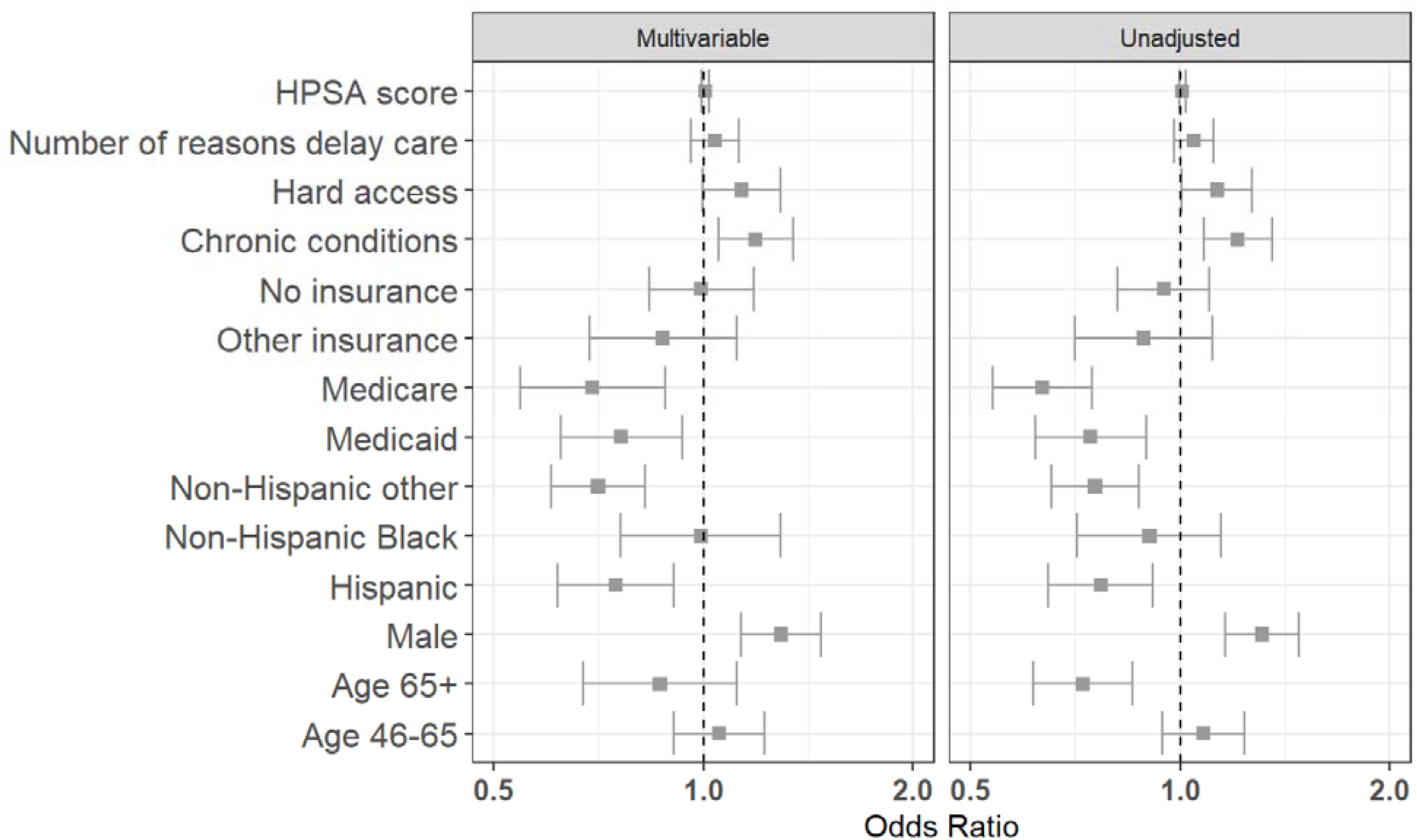
Prescribed medication.

**Figure 5b:**
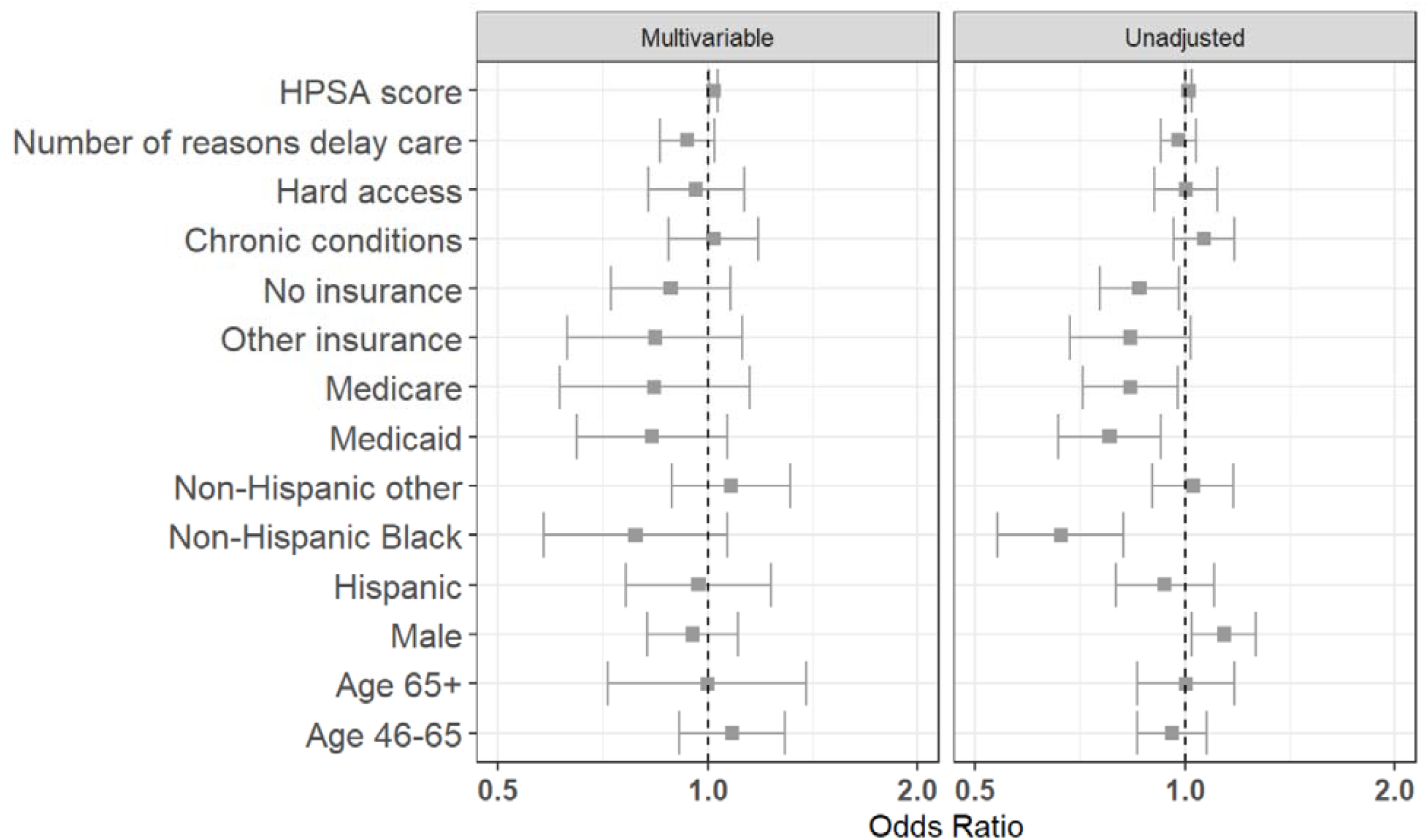
Prescribed medication within 1 day of symptom onset.

**Figure 5c:**
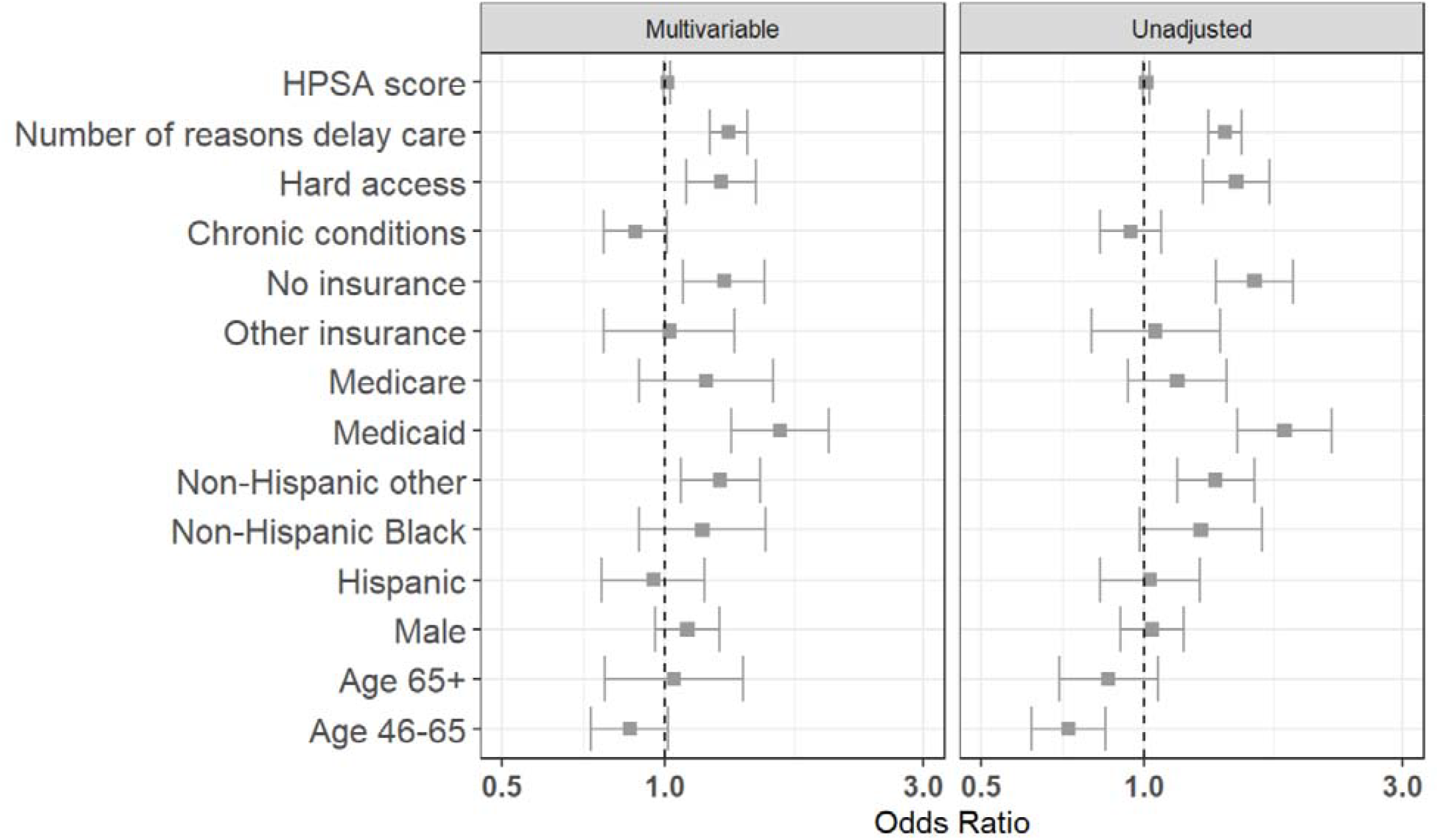
Opted for home-delivery of medication.

## Discussion

The Home Test-to-Treat (HTTT) program achieved high engagement and rapid antiviral treatment across a large, diverse population of adults. Over 83% of enrollees who tested positive for COVID-19 or flu proceeded to use the program’s telehealth service, and more than 80% of those were prescribed guideline-recommended antivirals. Consequently, a majority of treated patients began antiviral therapy promptly after symptom onset: 59.8% started medication within one day of symptom onset and over 92% within five days. Most telehealth consultations took place during evenings or weekends (less than a quarter occurred during weekday business hours), underscoring that HTTT successfully delivered care at times when traditional clinics are often unavailable. Together, these results demonstrate that a decentralized, at-home test-to-treat model can achieve prompt, widespread uptake of telehealth and same-day treatment initiation in practice.

The HTTT program’s treatment reach far exceeded what has been observed in traditional healthcare settings. Only about 24% of high-risk Veterans with COVID-19 infections received antiviral treatment.^1,2^ Other studies similarly reported antiviral uptake well under 35% in eligible patients.^3^ In contrast, HTTT prescribed oral antivirals to over 80% of infected enrollees, illustrating the impact of integrating services. The self-selected enrollees likely represent a more activated group of individuals. However, a study among predominantly vaccinated and activated patients, found that less than 20% of individuals who test positive received oral antivirals. Thus, the program’s one-stop approach: combining at-home testing, on-demand telehealth prescribing, and pharmacy or home delivery likely minimized the drop-off that occurs at multiple steps in usual care and increased adherence to guideline directed medical therapy as well as sped up the care cascade. This is critical because early antiviral therapy dramatically improves outcomes: for example, starting medication within 24–48 hours of symptoms can substantially reduce the risk of severe illness. Indeed, HTTT often enabled same-day diagnosis and treatment, an achievement that likely prevented a number of hospitalizations or complications.

A major finding is that HTTT provided equitable care on many key metrics. We observed no significant differences by race/ethnicity, insurance status, or prior access to care in the likelihood of receiving timely treatment once enrolled. By removing cost and transportation barriers and offering services remotely, HTTT appears to have narrowed or eliminated many of these disparities. In fact, the program reached groups that often struggle with access: enrollees who reported difficulty accessing healthcare were more likely to use telehealth when they became positive (aOR 1.35), suggesting HTTT successfully engaged those with unmet needs. At the same time, some differences persisted, highlighting areas for improvement. Older adults and Medicare beneficiaries were somewhat less likely to initiate a telehealth visit when positive, reflecting known digital access and literacy challenges among seniors. Additionally, among those who did use telehealth, antiviral prescribing rates were slightly lower for Hispanic patients and Medicare-insured patients. These residual gaps could stem from language or communication barriers, hesitancy to accept treatment, or potential provider biases in virtual care. Importantly, because HTTT removed many structural obstacles (free tests, free telehealth, no travel needed), the remaining disparities likely point to nuanced challenges such as technology comfort and cultural concordance. To fully close the remaining gaps, future programs could incorporate enhancements: for example, providing digital literacy support or phone-based options for older adults, and offering culturally and linguistically tailored outreach during telehealth encounters for minority patients. Such targeted improvements would build on HTTT’s strengths so that no subgroup is left behind in the test-to-treat process.

HTTT provides a proof of concept for decentralized care when the diagnostic and treatment cascade is clear, time sensitive, and protocolized. COVID-19 and influenza are appropriate use cases because accurate home tests are widely available, eligibility criteria for antivirals are standardized, and benefit depends on rapid initiation. Within this context, HTTT linked at-home testing, on-demand telehealth assessment, and swift medication fulfillment, achieving high telehealth uptake and timely prescribing at national scale, often outside business hours. Across many demographic categories the program delivered comparable timeliness once patients engaged the service, consistent with the view that structural access interventions can reduce inequities that persist in routine care. To translate this operational success to the broader crisis in American healthcare, similar services should be integrated with, rather than isolated from, existing systems through bidirectional exchange with EHRs and health information exchanges, automatic notification to usual clinicians, shared escalation pathways, closed-loop pharmacy fulfillment, and common quality and safety measures.

This study also had several limitations. First, as enrollees self-selected into the HTTT program (through convenience sampling), they likely differ from the general population in ways that could overestimate the feasibility and success of home-based interventions. Enrollees may have better digital access or health motivation than the general population. Second, key timeline data (symptom onset and test dates) were self-reported, introducing potential recall bias. This limits the accuracy of testing and treatment time periods. Third, the program was delivered via a single telehealth platform and pharmacy system, which may limit generalizability to other settings with different technologies or workflows. Fourth, we lacked detailed data on individuals who enrolled but did not utilize telehealth or did not report any positive test. Thus, we could not fully examine why some enrollees did not progress through the test-to-treat pathway (such as asymptomatic courses, competing priorities, or distrust).

In summary, HTTT offers an operational blueprint for rapid, equitable treatment at scale. However, this opportunity must be clear-eyed in the the objective to strengthen primary care capacity: routing straightforward, time-sensitive episodes through a coordinated decentralized pathway can free primary care teams to focus on longitudinal management and diagnostic complexity while extending after-hours coverage. Payment approaches should recognize triage, coordination, and supervisory time so that health systems and clinicians have aligned incentives to participate and to lead. If this model is extended beyond acute viral illness to chronic disease management or more intensive diagnostic assessment and titration, pairing the decentralized platform with a local, trusted workforce will be essential. Community health workers, pharmacists, nurses, and paramedicine teams can support home-based examination, specimen collection, imaging logistics, medication adjustment, culturally concordant counseling, and reliable follow-up, with clear handoffs, shared care plans, and safety nets for deterioration, as well as routine surveillance for adverse events and total cost. Given anticipated growth in Medicaid enrollment and the uninsured, this approach is actionable at federal, state, and county levels; public purchasers and health departments can procure integrated test-to-treat services, align them with FQHC and safety-net networks, and minimize patient out-of-pocket costs. The Rural Health Transformation Program can provide scaffolding for demonstration projects that adapt decentralized care to rural settings by supporting cross-sector partnerships, digital and broadband infrastructure, workforce development, and evaluation of integration, safety, equity, cost, and interoperability. A pragmatic agenda to build on this model across additional acute and chronic conditions, compare performance and timeliness with usual care, standardize minimum data elements and referral pathways, and identify which functions should remain local versus centralized is desperately needed.

## Notes

The authors thank the numerous partners of the Home Test-to-Treat program whose contributions were critical to the successful implementation of the program but did not rise to the level of co-authorship for the evaluation manuscript. In particular, we acknowledge Michael Wolfson, PhD (NIH/NIBIB) and Bruce J. Tromberg, PhD (NIH/NIBIB) for conceptual support and program guidance; Eric Gogstad (CDC) for expertise in program design and providing influenza antivirals from CDC; Dina Passman, PhD, MPH, Lisa Tung, PharmD, and Chris Crabtree, DrPH, MPH (ASPR) for conceptual design and implementation support, and providing Covid-19 tests and Covid-10 antivirals from ASPR; Michael Mina, MD, PhD (eMed) and the eMed team for modifying the digital platform and implementing the care model; VentureWell for primary program coordination and contracting. and the community engagement teams contracted by VentureWell for coordinating outreach to over 5,000 community organizations and programmatic support by Alex Guardo, Megan Aanstoos, and Rebekah Neal at VentureWell. We also thank all the telehealth clinicians, pharmacists, and staff who made the HTTT program possible. Most of all, we are grateful to the HTTT program enrollees for their trust and participation.

### Competing Interest Statement

The authors have declared no competing interest.

### Author Declarations

The study received full ethical approval from the Western Copernicus Group Institutional Review Board (Protocol # 1354231).

### Summary of Updates

Corrected acknowledgement section to rectify omission in attributions

